# *A MUC5B* gene polymorphism, rs35705950-T, confers protective effects in COVID-19 infection

**DOI:** 10.1101/2021.09.28.21263911

**Authors:** Anurag Verma, Jessica Minnier, Jennifer E Huffman, Emily S Wan, Lina Gao, Jacob Joseph, Yuk-Lam Ho, Wen-Chih Wu, Kelly Cho, Bryan R Gorman, Nallakkandi Rajeevan, Saiju Pyarajan, Helene Garcon, James B Meigs, Yan V Sun, Peter D Reaven, John E McGeary, Ayako Suzuki, Joel Gelernter, Julie A Lynch, Jeffrey M Petersen, Seyedeh Maryam Zekavat, Pradeep Natarajan, Cecelia J Madison, Sharvari Dalal, Darshana N Jhala, Mehrdad Arjomandi, Elise Gatsby, Kristine E Lynch, Robert A Bonomo, Mat Freiberg, Gita A Pathak, Jin J Zhou, Curtis J Donskey, Ravi K Madduri, Quinn S Wells, Rose DL Huang, Renato Polimanti, Kyong-Mi Chang, Katherine P Liao, Philip S Tsao, Peter W.F. Wilson, Adriana Hung, Christopher J O’Donnell, John M Gaziano, Richard L Hauger, Sudha K. Iyengar, Shiuh-Wen Luoh

## Abstract

**Rationale:** A common *MUC5B* gene polymorphism, rs35705950-T, is associated with idiopathic pulmonary fibrosis, but its role in the SARS-CoV-2 infection and disease severity is unclear.

**Objectives:** To assess whether rs35705950-T confers differential risk for clinical outcomes associated with COVID-19 infection among participants in the Million Veteran Program (MVP) and COVID-19 Host Genetics Initiative (HGI).

**Methods:** MVP participants were examined for an association between the incidence or severity of COVID-19 and the presence of a *MUC5B* rs35705950-T allele. Comorbidities and clinical events were extracted from the electronic health records (EHR). The analysis was performed within each ancestry group in the MVP, adjusting for sex, age, age^2,^ and first twenty principal components followed by a trans-ethnic meta-analysis. We then pursued replication and performed a meta-analysis with the trans-ethnic summary statistics from the HGI. A phenome-wide association study (PheWAS) of the rs35705950-T was conducted to explore associated pathophysiologic conditions.

**Measurements and Main Results:** A COVID-19 severity scale was modified from the World Health Organization criteria, and phenotypes derived from the International Classification of Disease-9/10 were extracted from EHR. Presence of rs35705950-T was associated with fewer hospitalizations (N_cases_=25353, N_controls_=631,024; OR=0.86 [0.80-0.93], p=7.4 × 10^−5^) in trans-ethnic meta-analysis within MVP and joint meta-analyses with the HGI (N=1641311; OR=0.89 [0.85-0.93], p =1.9 × 10^−6^). Moreover, individuals of European Ancestry with at least one copy of rs35705950-T had fewer post-COVID-19 pneumonia events (OR=0.85 [0.76-0.96], p =0.008). PheWAS exclusively revealed pulmonary involvement.

**Conclusions:** The *MUC5B* variant rs35705950-T is protective in COVID-19 infection.

## Introduction

A respiratory disease caused by a novel coronavirus was first reported towards the end of 2019, now known as SARS-CoV-2 (COVID-19). Despite massive public health measures and vaccination initiatives, the COVID-19 pandemic remains a major global health threat. By September 2, 2021, the coronavirus disease-2019 (COVID-19) pandemic had caused more than 219 million confirmed infections and more than 4.5 million deaths worldwide(1).

Parenchymal fibrosis is a late complication of respiratory infections with COVID-19(2–4). Among chronic lung diseases, idiopathic pulmonary fibrosis (IPF)(5), a disorder characterized by progressive pulmonary scarring which is associated with a median survival of 2-3 years in the absence of lung transplantation(6), shares several risk factors with those for severe COVID-19 disease, including advanced age(7), cardiovascular disease, diabetes, and history of smoking(5). Thus, common pathological processes may be shared between the fibrotic response towards COVID-19 infection and those underlying IPF.

IPF likely develops from a multifaceted interaction between genetic and environmental factors, age-related mechanisms, and epigenetic profibrotic reprogramming(8, 9). One of the most robust genetic risk factors identified for IPF susceptibility is rs35705950-T, a common G to T transversion located approximately 3 kb upstream of the mucin 5B, oligomeric mucus/gel-forming *MUC5B* gene (10, 11). Laboratory evidence supports that rs35705950-T is: 1) a functional variant located within an enhancer subject to epigenetic programming and 2) contributes to pathologic mis-expression in IPF (12).

Given the high minor allele frequency (MAF) of rs35705950-T (∼20% among individuals of European ancestry) and possible shared pathophysiological pathways between IPF and severe COVID-19 disease, we examined the association between rs35705950-T and the clinical outcomes of COVID-19 infection in the Million Veteran Program (MVP), a multi-ethnic cohort of over 650,000 U.S. Veterans with detailed EHR and genotyping data(13). Following our primary analysis in the MVP, we validated our results with a comparable analysis conducted in the Host Genetics Initiative (HGI), a global collaboration of over 160 genetic studies assembled to facilitate rapid discovery and dissemination of COVID-19 related science (14).

## Methods

### Data Sources

Data from the MVP, a multi-ethnic genetic biobank sponsored by the United States Veterans Affairs (VA), were analyzed (13). All protocols were approved by the VA Central Institutional Review Board and all participants provided written informed consent. For detailed Materials and Methods, please see **methods in the online data supplement.**

Demographic and pre-existing comorbidity data were collected from questionnaires and the VA EHR; “pre-COVID” data was from the time of enrollment into the MVP to September 30, 2019. The cohort demographics and a description of the clinical conditions for all tested patients in the two years preceding the index dates are presented in a supplemental table **(Table E1)**.

Genotyping was performed using a custom Thermo Fisher Axiom genotyping platform (MVP 1.0) which included direct genotyping of rs35705950-T. Ancestry was defined using Harmonized ancestry, race, and ethnicity (HARE) derived from self-report and genetic ancestry data(15).

#### COVID-19 outcome definitions

COVID-19 infection status from 02/2020 - 04/2021 was assessed by either self-report (if testing was performed outside the VA) or by a positive polymerase chain reaction (PCR)-based test(16, 17). For subgroup analyses of severity, only patients with confirmed PCR-based tests were examined. The *index date* was defined as a COVID-19 diagnosis date, i.e., specimen date, or a self-reported date of diagnosis; and for a hospitalized patient, the admission date up to 15 days prior to the COVID-19 case date.

Our analyses used harmonized definitions with the HGI to enable us to obtain larger sample sizes and consistent results. In accordance with the HGI definitions, the three following analyses were performed: (1) COVID Susceptibility: COVID-19 positive vs. population controls; (2) COVID Hospitalization-v1: COVID-19 positive and hospitalized vs. population controls; (3) COVID-19 hospitalization-v2: COVID-19 positive and hospitalized vs. COVID-19 positive but not hospitalized.

Our other analyses focused on data present in MVP only and addressed the outcome severity and post-index events. For these sets of analyses, we only focused on patients who received their PCR-based COVID-19 testing within VA systems. COVID-19 severity scale was derived from the WHO COVID-19 Disease Progression Scale(18) as mild, moderate (hospitalization), severe (Intensive Care Unit-level care), or death within 30 days of PCR-confirmed COVID-19 infection. All data and variables were assessed centrally by the MVP data core’s Shared Data Repository (SDR).

#### Post-index analytic constructs and study design

The ICD codes used to pull the pneumonia events within 60 days post-index (pneumonia60d) are presented in **Table E2**. Pre-index conditions were derived using natural language processing (NLP)-boosted unstructured notes, ICD and Current Procedural Terminology (CPT) codes, and medications are taken 2 years prior. Post-index conditions, including pneumonia, were derived using ICD and CPT codes, and medications 60 days after the index date. Association with post-index pneumonia events (pneumonia60d) were performed among patients who received confirmatory COVID-19 PCR testing at VA sites.

### Statistical analysis

Firth logistic regression(19, 20) as implemented in the R (v3.6.1) package “brglm2” (version 0.7.1)(21) was used to examine the association between COVID-19 outcomes and rs35705950-T (additive model) separately by ancestry, with adjustment for age, age^2^, sex, and ethnicity-specific principal components. Trans-ethnic meta-analyses were performed using random-effects models in “metafor” (version 2.4-0)(22). Interactions between COVID-19 infection status and rs35705950-T on the outcome of post-index pneumonia at 60 days were assessed using a multiplicative interaction term followed by stratified analyses by COVID-19 infection status, with additional covariate adjustment of pre-index pneumonia.

### Phenome-wide and Laboratory-wide association studies (PheWAS and LabWAS)

Associations between rs35705950-T allele and pre-existing comorbid conditions and laboratory values were examined using preclinical data prior to the COVID-19 era (Sept 2019). Individuals with ≥2 Phecodes(23) were defined as cases. Phecodes with <200 cases within each ancestry group were excluded, resulting in 1618 (EUR), 1289 (AFR), 994 (HIS) Phecodes. LabWAS was conducted for 69 clinical tests; for individuals with repeated measures, the median of the individuals’ EHR record was used. Logistic/Firth regression and linear regression were used for Phecodes and laboratory measurements, respectively. Bonferroni-adjusted thresholds for significance (by ancestry) were: EUR = 3.09 × 10^−05^ (0.05/1618), AFR = 3.8 × 10^−05^ (0.05/1289), HIS = 5.03 × 10^−05^(0.05/994). Analyses were performed using PLINK2(24) **(Additional details in supplemental methods)**.

### Meta-analysis with HGI

Data from Release 5 (01/18/2021) of the COVID-19 Host Genetics Initiative (HGI) were utilized for replication via an inverse-variance weighted meta-analysis using plink2a(24) and GWAMA(25)**(Additional details in supplemental methods)**.

## Results

### Elucidation of the shared genetics with the MUC5B rs35705950-T by PheWAS and LabWAS

In order to understand the pathophysiology associated with the *MUC5B* rs35705950-T allele, and more specifically how the presence of the *MUC5B* rs35705950-T allele(s) might impact the susceptibility and severity of COVID-19, we performed PheWAS and LabWAS to search for the *MUC5B* rs35705950-T allele associated conditions prior to COVID-19 infection. The sample sizes for MVP participants used for PheWAS and COVID-19 association studies, as well as HGI participants examined in this study, are shown in **Table 1 (Figure E1)**. The results of the PheWAS are shown in **Figure 1 and Table E3**.

**Table 1.**
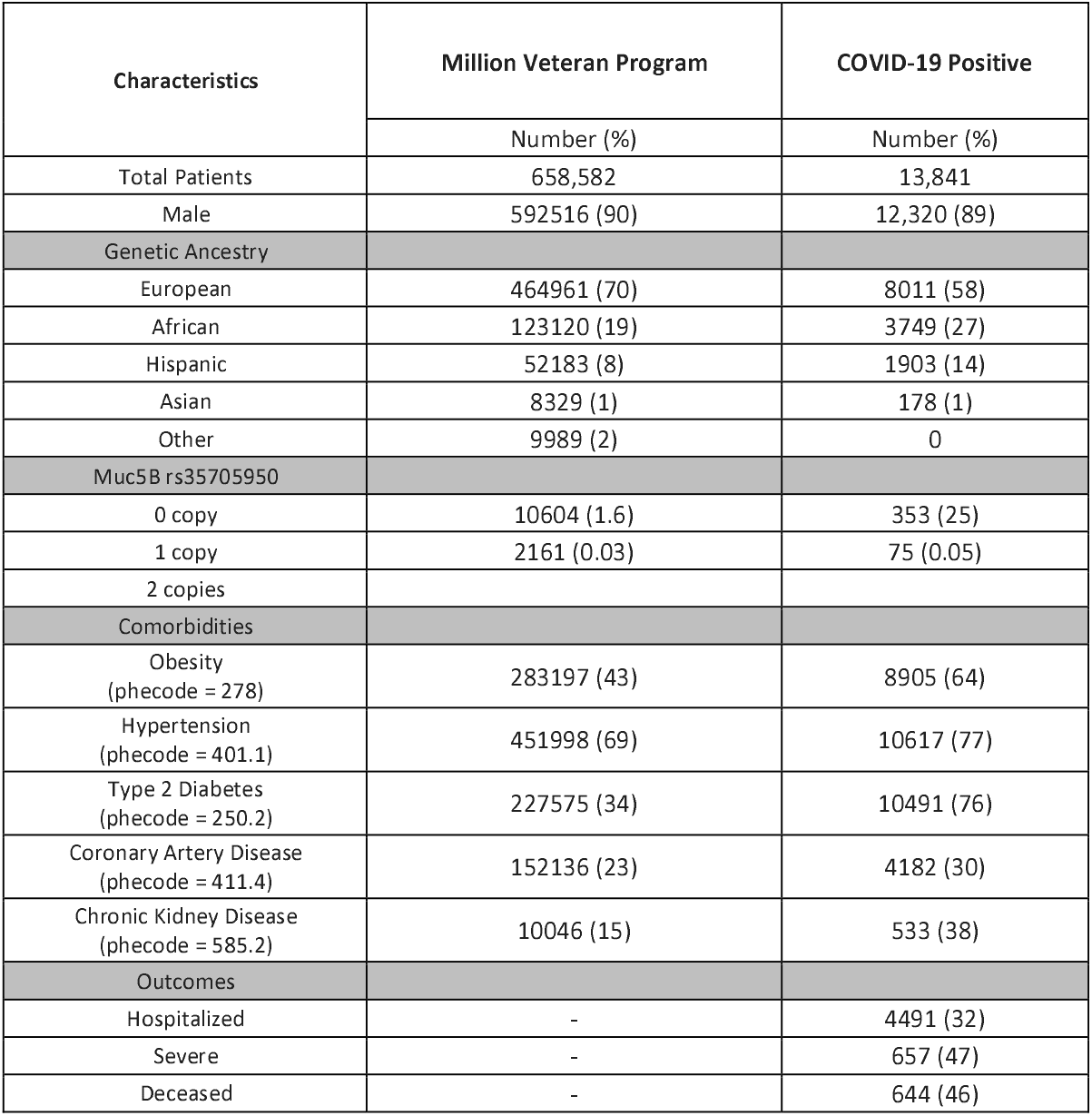
Demographics for COVID-19 tested positive and all MVP participants examined in this study.

**Figure 1.**
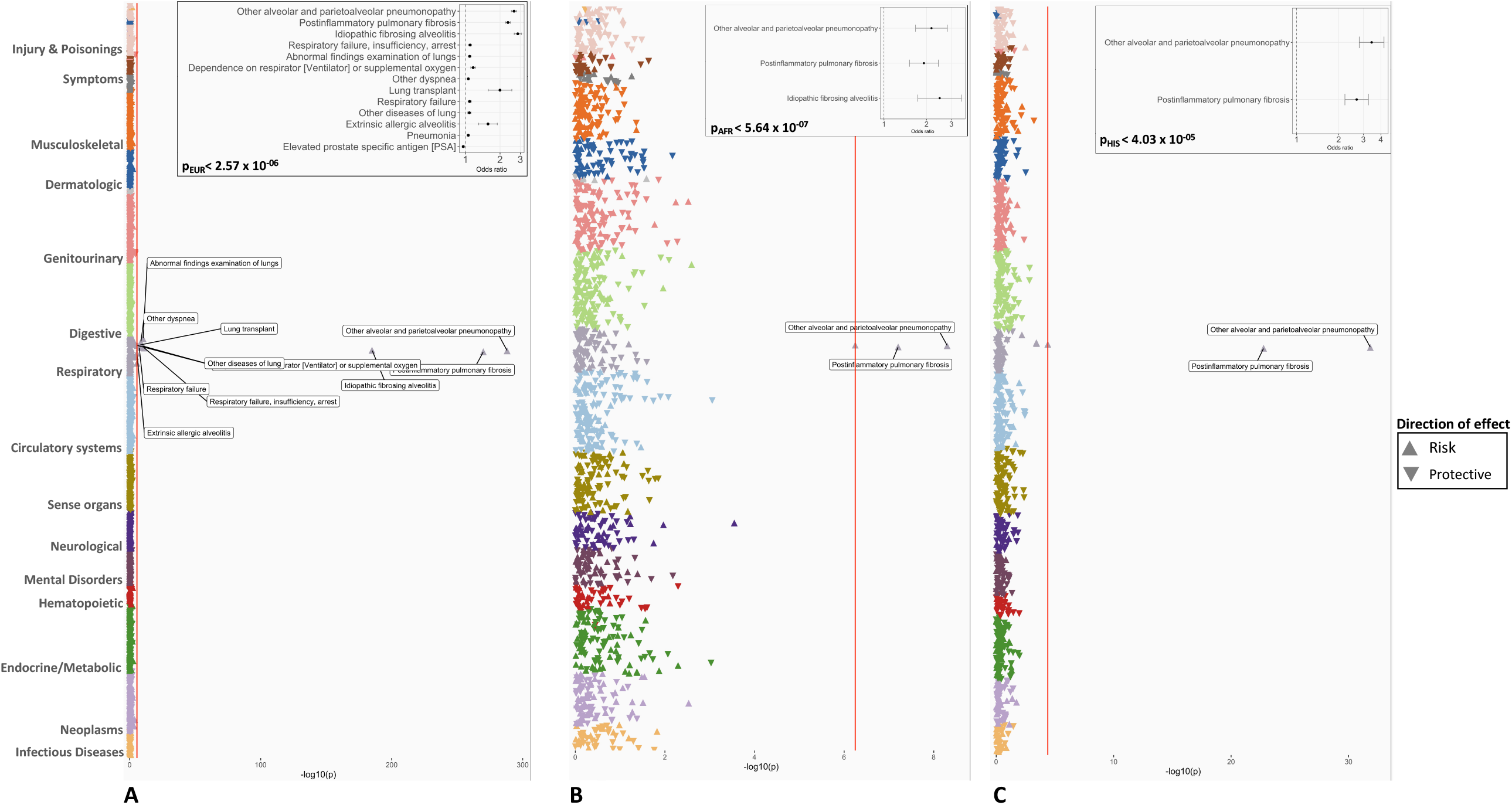
Phenome-Wide Association Study (PheWAS) of *MUC5B* rs35705950 allele in the Million Veteran Program. A PheWAS plot shows associations of rs35705950 and phenotypes derived from the electronic health records data prior to COVID-19 in MVP participants from A) European ancestry B) African ancestry and C) Hispanic ancestry. The phenotypes are shown on the x-axis and organized by disease categories. The p-value (-log10) of each association is shown on the y-axis the direction of the triangle represents the direction of effect of the associations - with the upward triangle as increased risk and the downward triangle as reduced risk. The red line indicates the significance threshold based on the Bonferroni correction. The forest plot of Bonferroni significant associations are shown within the right top corner of each PheWAS plot. The Bonferroni threshold for each ancestry group is shown in the forest plot.

In the PheWAS analysis between this *MUC5B* variant and 1605 phenotypes (cases > 200) from participants of European ancestry, we found significant associations (P_bonferroni_ < 2.5 × 10^−6^) with 12 respiratory conditions. Consistent with the previous finding in IPF, rs35705950-T was associated with increased risk of Idiopathic fibrosing alveolitis (phecode = 504.1; OR = 2.85 [2.65 -3.05], P = 8.90 × 10^−186^), other alveolar and parietoalveolar pneumonopathy (phecode = 504; OR = 2.64 [2.50 - 2.78], P = 7.07 × 10^−289^), and postinflammatory pulmonary fibrosis (phecode = 502; OR = 2.34 [2.23 - 2.45], P = 8.90 × 10^−186^). Additionally, we also observed significant associations with respiratory failure (Phecode: 509), ventilatory dependence (Phecode 509.8), lung transplant (Phecode: 510.2) and pneumonia (Phecode: 480) **(Figure 1, Table E3)**. Notably, we evaluated Phecodes for influenza infection (481) in our PheWAS analysis and did not observe an association with *MUC5B* rs35705950-T (p<0.05; the power to detect a difference was >95% as there were 4728 cases of influenza in EUR).

We identified, as in EUR, a significant association of this *MUC5B* variant with an increased risk of three pulmonary conditions in African ancestry participants: idiopathic alveolitis (Phecode: 504.1), other alveolar and parietoalveolar pneumonopathy (Phecode:504), and post-inflammatory fibrosis (Phecode: 502) **(Figure 1, Table E3)**. Two of these associations, other alveolar and parietoalveolar pneumonopathy (Phecode:504) and post-inflammatory fibrosis (Phecode: 502), were also seen in HIS ancestry, suggesting shared etiology.

We performed a Laboratory-wide association study of the *MUC5B* rs35705950-T with median values of clinical laboratory tests measured prior to the COVID-19 pandemic. We only included quantitative traits with 1000 or more individuals. Among EUR participants, we evaluated 63 lab measurements and 10 had a significant association with the rs35705950-T. Increased level of neutrophils (absolute count) had the most significant association (beta= 0.05, p=6.24 × 10^−23^). This specific association has not been previously reported. Other significant associations with increased levels were white blood cell counts, neutrophil fraction, estimated glomerular filtration rate (eGFR), eosinophils (absolute count), monocytes (absolute count), and platelets **(Figure 2, Table E4)**. The variant had an association with reduced levels of albumin, lymphocyte fraction, and creatinine **(Figure 2, Table E4)**. There was no significant association with lab measurements in AFR or HIS, but among HIS monocytes (absolute count) were significant (beta =0.0078, p 1.66 × 10^−04^) in the same direction as in EUR.

**Figure 2.**
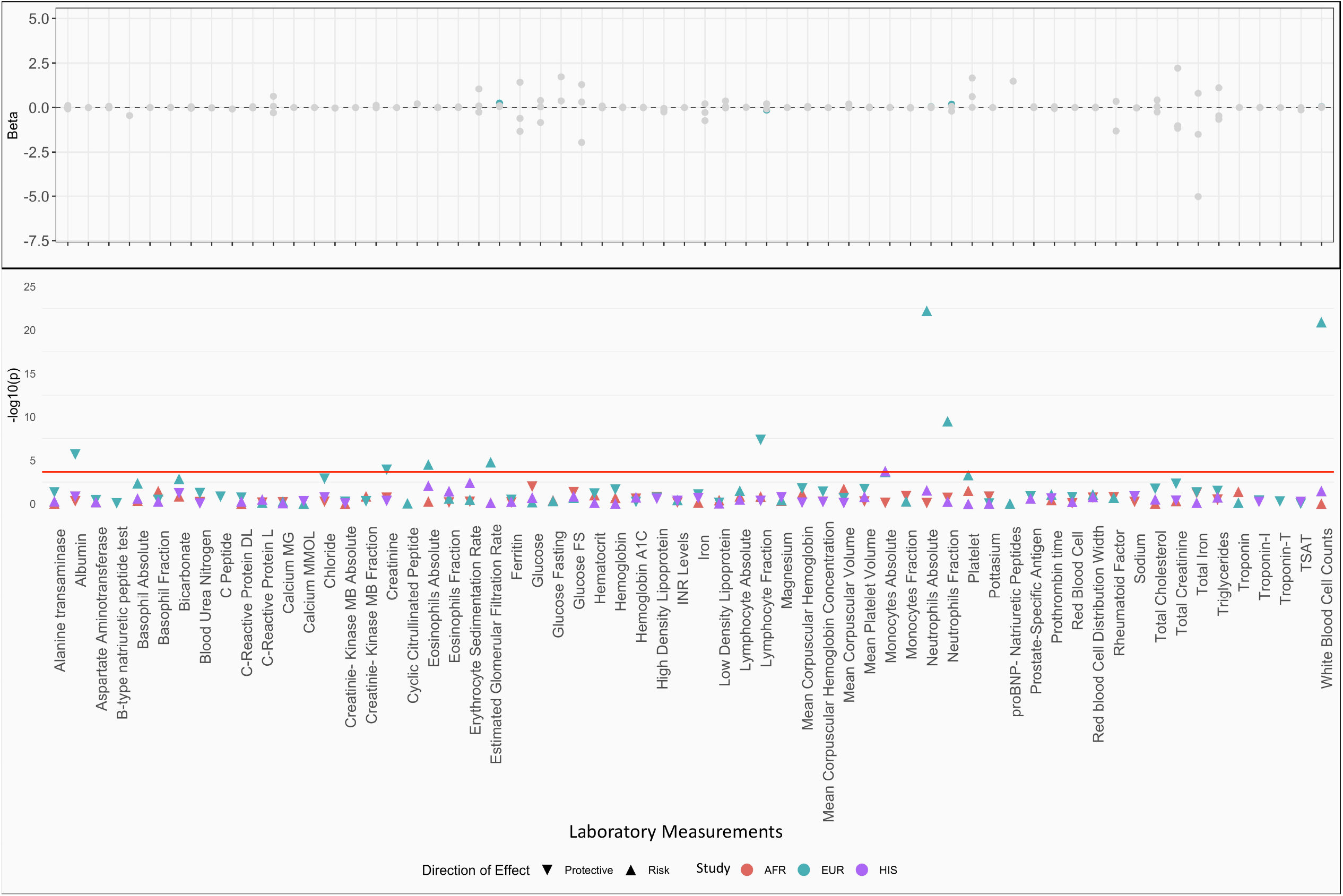
Laboratory-Wide Association Study (PheWAS) of *MUC5B* rs35705950 allele in the Million Veteran Program. A LabWAS plot shows associations of rs35705950 and median values of laboratory measures extracted from electronic health records data prior to COVID-19 in MVP participants. The bottom panel shows the -log10 (p-value) on the y-axis and laboratory test descriptions on the x-axis. Triangles points up have increasing effects and points down have decreasing effects. The colors represent the different ancestry groups. The top panel shows beta from the regression model for each laboratory measure. The significant results are highlighted in the color corresponding to ancestry groups and other results are plotted in grey.

### Association of the *MUC5B* rs35705950-T allele with the COVID-19 infection or hospitalization in the MVP and meta-analysis with HGI

We tested for association between *MUC5B* rs35705950-T with three COVID-19 phenotype definitions 1) COVID-19 positive as cases vs all the other participants in the MVP as controls 2) COVID-19 positive that required hospitalization for treatment vs all the other participants in the MVP as controls 3) COVID-19 positive that required hospitalization for treatment vs COVID-19 positives that didn’t require hospitalization as controls. First, we performed the analysis in three major ancestries separately (European, African, and Hispanic). Then, we meta-analyzed the summary statistics with the COVID-19 HGI (Freeze 5) using an inverse-variance weighted method (GWAMA)(25). Among the three COVID-19 phenotypes, the most significant association of rs35705950-T allele carriers was with fewer hospitalization events (OR = 0.89 [0.85-0.93], p=1.88 × 10^−6^, **Figure 3** and **Table 2**).

**Table 2.**
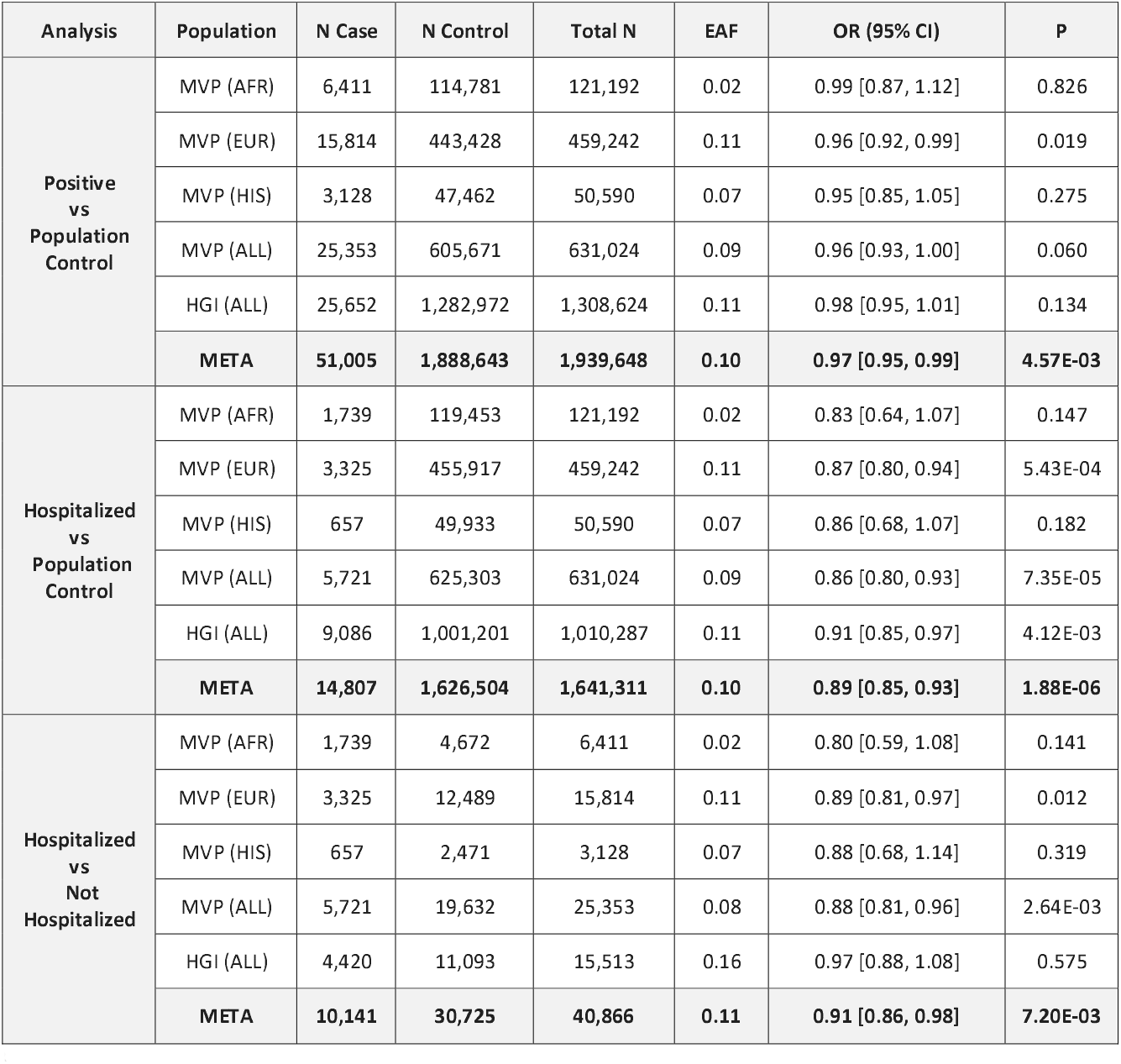
Association of rs35705950 in *MUC5B* with (i) COVID-19 Positive vs Population Controls, (ii) COVID-19 Positive, Hospitalized vs Population Controls, and (iii) COVID-19 Positive, Hospitalized vs COVID-19 Positive, not Hospitalized. Odds ratio (OR) and 95% confidence interval (95% CI) is reported for the minor (T) allele, and results are shown for VA Million Veteran Program (MVP) African Americans (AFR), European Americans (EUR), Hispanic/Latino Americans (HIS), and trans-ethnic meta-analysis (ALL), the COVID-19 Host Genetics Initiative (HGI) trans-ethnic release 5 meta-analysis excluding MVP and 23&Me, and the meta-analysis of MVP and HGI (META).

**Figure 3.**
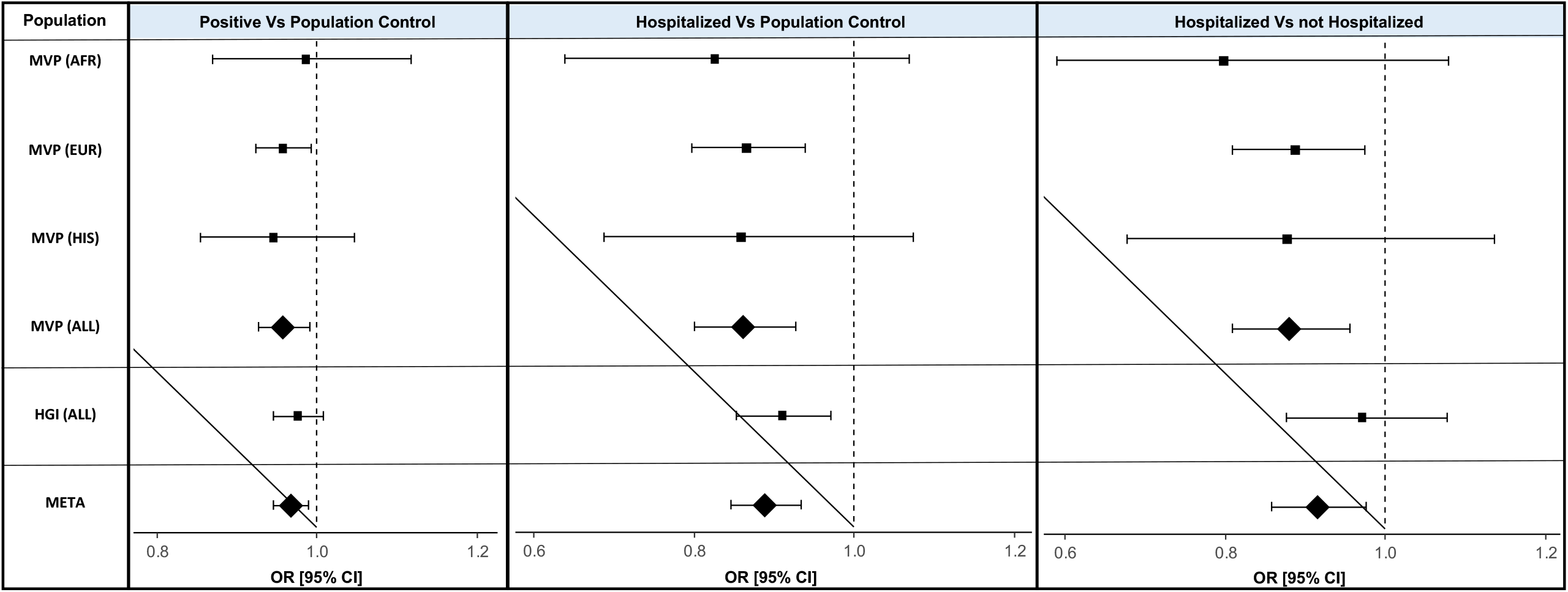
Forest plot association of rs35705950 in *MUC5B* with (i) COVID-19 Positive vs Population Controls, (ii) COVID-19 Positive, Hospitalized vs Population Controls, and (iii) COVID-19 Positive, Hospitalized vs COVID-19 Positive, not Hospitalized. Odds ratio (OR) and 95% confidence interval (95% CI) is reported for the minor (T) allele, and results are shown for VA Million Veteran Program (MVP) African Americans (AFR), European Americans (EUR), Hispanic/Latino Americans (HIS), and trans-ethnic meta-analysis (ALL), the COVID-19 Host Genetics Initiative (HGI) trans-ethnic release 5 meta-analysis excluding MVP and 23&Me, and the meta-analysis of MVP and HGI (META).

### Association of the *MUC5B* rs35705950-T allele with fewer pneumonia events within 60 days of COVID-19 infection in the MVP

In 9,216 COVID-19 infected MVP patients, the adjusted odds ratio for post-index pneumonia was 14.8% less with each additional *MUC5B* rs35705950-T allele (OR = 0.852 [0.757-0.958], p=0.008). In COVID-19 negative patients, the adjusted odds for post-index pneumonia was 7.8% higher with each additional *MUC5B* rs35705950-T allele (OR=1.078 [1.001-1.162], p=0.048). This differential effect of an additional *MUC5B* rs35705950-T allele on post-index pneumonia in COVID-19 positive vs. COVID-19 negative patients was statistically significant (p-value for interaction 0.0009) in EUR (**Table 3, Table E5**).

**Table 3.** Fewer pneumonia events developed within 60 days post COVID-19 infection for MVP EUR individuals with the presence of a *MUC5B* rs35705950 allele. Odds ratios are estimated from Firth logistic regression adjusting for pre-index pneumonia, age, age^2^, and PC1-20, including an interaction between additive MUC5B rs35705950 allele and COVID-19 infection.

|  | COVID-19 negative | COVID-19 positive | COVID-19 & *MUC5B*<br>p-value for interaction |
| --- | --- | --- | --- |
|  | OR (95% CI) of a *MUC5B* allele |  | p=0.0009 |
|  | 1.08 (1.00, 1.16)<br>p=0.048 | 0.89 (0.76, 0.96)<br>p=0.008 |  |
| *MUC5B*=0 | *MUC5B*=1 | *MUC5B*=2 |  |
| OR (95% CI) of COVID-19 positive status |  |  |  |
| 10.00 (9.35, 10.70)<br>p<0.0001 | 7.91 (6.97, 8.97)<br>p<0.0001 | 6.26 (4.83, 8.08)<br>p<0.0001 |  |

### Association of the *MUC5B* rs35705950-T allele with severe outcomes of COVID-19 infection in the MVP

Presence of a *MUC5B* rs35705950-T allele was not associated with severe outcomes of COVID-19 infection in the MVP. The *MUC5B* rs35705950-T allele was not associated with severe outcomes with mortality (OR = 1.01 [0.58-1.20], p= 0.72) nor mortality alone (OR = 0.91 [0.72-1.16], p=0.25) in EUR ancestry individuals(**Table E6**).

## Discussion

The data herein establishes that the “T” allele of rs35705950-T in *MUC5B*, which has been associated with an *increased* risk for the development of IPF, confers a *decreased* risk of hospitalization and pneumonia following COVID-19 infection among MVP participants of European ancestry. The protective effect of the rs35705950-T, in addition to being counterintuitive, is in stark contrast to the increased risk of severe COVID-19 disease observed for other well-established causal variants or IPF, including variants located in the *TERC, DEPTOR*, and *FAM13A(26)*.

The protein product of *MUC5B* is a major gel-forming mucin in the lung that plays a key role in mucociliary clearance (MCC) and host defense(27). MUC5B protein is secreted from proximal submucosal glands and distal airway secretory cells(28–30). Mucus traps inhaled particles, including bacteria, and transporting them out of the airways by ciliary and cough-driven forces. Mucin also helps remove endogenous debris including dying epithelial cells and leukocytes. MUC5AC and MUC5B are two major secreted forms of mucins in the lung.

The rs35705950-T is located within an enhancer region of *MUC5B*; the “T” allele demonstrates gain-of-function and is associated with enhanced expression of the *MUC5B* transcript in lung tissue from unaffected subjects and patients with IPF(31). In patients with IPF, excess MUC5B protein is especially observed in epithelial cells in the respiratory bronchiole and honeycomb cyst(29, 30, 32), regions of the lung involved in lung fibrosis.

Mouse models found that *Muc5b* is required for mucociliary clearance, for controlling bacterial infections in the airways and middle ear, and for maintaining immune homeostasis in mouse lungs(33). Muc5b deficiency caused materials to accumulate in the upper and lower airways. This defect led to chronic infection by multiple bacterial species, including *Staphylococcus aureus*, and to inflammation that failed to resolve normally. Apoptotic macrophages accumulated, phagocytosis was impaired, and interleukin-23 (IL-23) production was reduced in *Muc5b*(-/-) mice. By contrast, in transgenic mice that overexpress *Muc5b*, macrophage functions improved (33). *Muc5B* over-expressing transgenic mice have been shown to be more susceptible to the fibroproliferative effects of bleomycin (34), consistent with a role in IPF. Paradoxically, while the “T” allele of rs35705950-T increases susceptibility towards the development of IPF, the same allele has also been associated with *decreased* mortality among IPF patients(35).

Our analyses demonstrating a significant interaction between COVID-19 infection and the prospective development of pneumonia suggest a possible mechanism by which the protective effect of rs35705950-T is mediated. Whether enhanced pulmonary macrophage function or quantitative or qualitative changes in mucous production resulting from the minor allele of rs35705950-T are responsible for the observed protective effect should be explored in future work. Of note, the *MUC5B* rs35705950-T allele did not decrease the risk of pneumonia in COVID-19 tested negative participants (**Table 3**), suggesting that the protective effect may be specific to COVID-19 related pneumonia. More studies in the future are needed to further investigate this phenomenon.

No extrapulmonary association was noted on PheWAS analysis suggesting a very circumscribed molecular and clinical effect of this promoter variant. This supports the notion that the effect of rs35705950-T on COVID-19 infection is mediated in pulmonary tissues. The *Muc5b* over-expression in the distal airway may specifically or non-specifically affect the SARS-CoV-2 viral infection in the lung, leading to decreased incidence of pneumonia and hospitalization in the infected individuals.

The human *MUC5B* rs35705950-T allele does not appear to be sufficient to cause pulmonary fibrosis. Although ∼20% of the non-Hispanic white populations have a copy of the *MUC5B* rs35705950-T allele(31, 33), IPF is a rare disease with a population prevalence of less than 0.1% (36). Additional genetic and/or environmental insults are likely needed in the development of IPF in humans. Since the overwhelming number of individuals with the *MUC5B* rs35705950-T allele will not know their *MUC5B* status, it is unlikely that the reason for our observation is due to a change in health behaviors of participants that carry this variant.

The *MUC5B* rs35705950-T allele was associated with elevated neutrophil counts. This could be due in part to the association of this allele with an increased incidence of pneumonia. It is worth noting that neutrophils are a major source of alpha-defensin and elevated alpha-defensin levels were seen in the serum of IPF patients; the levels of alpha-defensin in the serum correlated with the lung function decline in the IPF patients(37, 38).

Longer follow-up of SARS-CoV-2 infected individuals with the *MUC5B* rs35705950-T allele is needed. One would need to be cautious regarding the longer-term outcome of COVID-19 in the *MUC5B* rs35705950-T allele positive individuals as a fibrotic response has been reported in the survivors of severe COVID-19. This is of particular importance if the manipulation of *MUC5B* expression is considered in the prevention/treatment of COVID-19.

The *MUC5B* rs35705950-T allele variant resides within an enhancer subject to lineage- and disease-dependent epigenetic remodeling. It was postulated that this G to T transversion in the *MUC5B* rs35705950-T allele might lead to the removal of a binding site for the GCF transcription repressor(12, 39). A potential avenue for chromatin-based therapies in which *MUC5B* enhancer chromatin architecture serves as a target to block the *MUC5B* mis-expression was proposed(12, 39). Additional small molecule and signaling inhibitors targeting IPF are being studied as well(40). These strategies are generally aiming at reducing fibrosis or the effects associated with *MUC5B* over-expression. How these strategies or alternatives can be utilized to treat/prevent COVID-19 remains to be studied.

In conclusion, we show in this study a common *MUC5B* promoter variant leading to *MUC5B* over-expression is associated with fewer hospitalizations and pneumonia events after SARS-CoV-2 infection. Our study provides a strong rationale to stratify patient populations based on common and disease-related genetic polymorphism in order to better understand the mechanisms and their clinical implications in COVID-19. How the *MUC5B* rs35705950-T allele association may shed light on the pathogenesis and/or management of COVID-19 remains to be fully examined.

### Strengths & Limitations

MVP is a large genomic medicine database with diverse ethnicity and geography. MVP participants are predominantly males but it represents a large multi-ethnic, prospective cohort available. Successful replication in the HGI and meta-analysis is a strength as well as our ability to investigate specific clinical events post-index. PheWAS was designed as a broad screen to test for potentially clinically relevant associations between genes and phenotypes and helped in the understanding of potential disease mechanisms but has limited power to detect associations among uncommon conditions, especially when further stratified by genetic ancestry.

## Data Availability

Full summary-level association data from the PheWAS from this study is made available as supplementary files.

## Acknowledgments

This research is based on data from the Million Veteran Program, Office of Research and Development, Veterans Health Administration, and was supported by award MVP035. This publication does not represent the views of the Department of Veteran Affairs of the United States Government.

We are grateful to our Veterans for their contribution to MVP. Full acknowledgments for the VA Million Veteran Program COVID-19 Science Initiative can be found in the supplemental methods.

## Conflict of Interest

CJO is an employee of Novartis Institute for Biomedical Research. PN reports grant support from Amgen, Apple, AstraZeneca, Boston Scientific, and Novartis, personal fees from Apple, AstraZeneca, Blackstone Life Sciences, Genentech, and Novartis, and spousal employment at Vertex, all unrelated to the present work.

## Notes

Supported by MVP035 award from Million Veteran Program, Office of Research and Development, Veterans Health Administration, and Veteran Affairs of the United States Government BX 004831 (P.W/K.C). This publication does not represent the views of the Department of Veteran Affairs of the United States Government.

### Author Declarations

This research is based on data from the Million Veteran Program, Office of Research and Development, Veterans Health Administration, and was supported by award MVP035. All protocols were approved by the VA Central Institutional Review Board and all participants provided written informed consent.

