## Supplemental Text for "*A MUC5B* gene polymorphism, rs35705950-T, confers protective effects in COVID-19 infection"

^1^Corporal Michael Crescenz VA Medical Center, Philadelphia, PA, 19104, USA, ^2^Department of Medicine, Perelman School of Medicine, University of Pennsylvania, Philadelphia, PA, 19104, USA, ^3^OHSU-PSU School of Public Health, Oregon Health & Science University, Portland, OR, 97239, USA, ^4^Knight Cancer Institute, Biostatistics Shared Resource, Oregon Health & Science University, Portland, OR, 97239, USA, ^5^VA Portland Health Care System, OR, 97239, USA, ^6^MAVERIC, VA Boston Healthcare System, Boston, MA, 02130, USA, ^7^Department of Medicine, Pulmonary, Critical Care, Sleep, and Allergy Section, VA Boston Healthcare System, Boston, MA, 02115, USA, ^8^Channing Division of Network Medicine, Brigham & Women's Hospital, Boston, MA, 02115, USA, ^9^Knight Cancer Institute, Biostatistics Shared Resource, Oregon Health & Science University, ^10^VA Portland Health Care System, ^11^Department of Medicine, VA Boston Healthcare System, Boston, MA 02115, USA, ^12^Brigham & Women's Hospital, ^13^MAVERIC, VA Boston Healthcare System, Boston, MA, 2130, USA, ^14^Department of Medicine, Cardiology, Providence VA Healthcare System, Providence, RI, 02908, USA, ^15^Alpert Medical School & School of Public Health, Brown University, Providence, RI, 02903, USA, ^16^MAVERIC, VA Boston Healthcare System, 150 S Huntington Ave, Boston, MA, 02130, USA, ^17^Medicine, Aging, Brigham and Women's Hospital, Harvard Medical School, Boston, MA, 02130, USA, ^18^VA Boston Healthcare System, 150 S Huntington Ave, Boston, MA, 02131, USA, ^19^Yale Center for Medical Informatics, Yale School of Medicine, New Haven, CT, 06511', USA, ^20^Clinical Epidemiology Research Center (CERC), VA Connecticut Healthcare System, West Haven, CT, 06516', USA, ^21^VA Boston Healthcare System, Boston, MA, 2115, ^22^Department of Medicine, Harvard Medical School, ^23^Medicine, General Internal Medicine, Massachusetts General Hospital, 55 Fruit St, Boston, MA, 02115, USA, ^24^Epidemiology, Emory University School of Public Health, 1518 Clifton Rd. NE, Atlanta, GA, 30322, USA, ^25^Atlanta VA Health Care System, 1670 Clairmont Road, Decatur, GA, 30033, USA, ^26^Deptartment of Medicine, Phoenix VA Healthcare System, Phoenix, AZ, 85012, USA, ^27^Univ. of AZ, ^28^Department of Psychiatry and Human Behavior, Providence VA Medical Center, Providence, RI, 2908, USA, ^29^Brown University Medical School, ^30^Department of Medicine, Gastroenterology, Durham VA Medical Center, 508 Fulton St, Durham, NC, 27705, USA, ^31^Department of Medicine, Gastroenterology, Duke University, Durham, NC, 27710, USA, ^32^Psychiatry, Human Genetics, Yale Univ. School of Medicine, 950 Campbell Avenue, West Haven, CT, 06516, USA, ^33^VA CT Healthcare Center, ^34^VA Informatics & Computing Infrastructure, VA Salt Lake City Health Care System, Salt Lake City, UT, USA, ^35^Department of Medicine, University of Utah School of Medicine, Salt Lake City, UT, ^36^Pathology and Laboratory Medicine, Corporal Michael Crescenz VA Medical Center, Philadelphia, PA, 19104, USA, ^37^Perelman School of Medicine, University of Pennsylvania, Philadelphia, PA, 19104, USA, ^38^Computational Biology & Bioinformatics, Yale School of Medicine, 333 Cedar St, New Haven, CT, 6510, USA, ^39^Program in Medical and Population Genetics, Cardiovascular Disease Initiative, Broad Institute of Harvard and MIT, Cambridge, MA, 02142, USA, ^40^Cardiovascular Research Center, Massachusetts General Hospital, Boston, MA, 02114, USA, ^41^Department of Medicine, Harvard Medical School, Boston, MA, 02115, USA, ^42^Clinical Data Science Research Group, ORD, Portland VA Medical Center, Portland, OR, 97239, USA, ^43^Pathology and Laboratory Medicine, Corporal Michael J Crescenz VA Medical Center, Philadelphia, PA, 19104, USA, ^44^Pathology and Laboratory Medicine, Corporal Michael Crescenz VA Medical Center, Philadelphia, PA, 19104, ^45^Perelman School of Medicine, University of Pennsylvania, 19104, ^46^Medicine, Pulmonary and Critical Care, San Francisco VA Healthcare System; University of California San Francisco, San Francisco, CA, 94121, USA, ^47^VA Informatics and Computing Infrastructure (VINCI), VA Salt Lake City Healthcare System, Salt Lake City, UT, ^48^Internal Medicine, Epidemiology, University of Utah School of Medicine, Salt Lake City, UT, ^49^(No affiliation data provided), ^50^Department of Psychiatry, Division of Human Genetics, Yale School of Medicine, New Haven, CT, 06511, USA, ^51^VA Connecticut Healthcare System, West Haven, CT, 06516, USA, ^52^Medicine, University of California, Los Angeles, Los Angeles, CA, 90024, USA, ^53^Epidemiology and Biostatistics, University of Arizona, AZ, 85724, USA, ^54^Infectious Disease Section, Louis Stokes Cleveland VA and Case Western Reserve University, Cleveland, OH, 44106, USA, ^55^Data Science and Learning, Argonne National Laboratory, 9700 S Cass Ave, Lemont, IL, 60439, ^56^Departments of Medicine, Biomedical Informatics, and Pharmacology, Vanderbilt University Medical Center, Nashville, TN, 37232, USA, ^57^Corporal Michael J Crescenz VA Medical Center, ^58^Medicine, Rheumatology, VA Boston Healthcare System, Boston, MA, 02130, ^59^Precision Medicine, VA Palo Alto Health Care System, 3801 Miranda Avenue, Palo Alto, CA, 94304, USA, ^60^Emory University School of Medicine, Atlanta, GA, 30322, USA, ^61^Medicine, Cardiology, VA Boston Healthcare System, 1400 VFW Parkway, Boston, MA, 2132, USA, ^62^VA Boston Healthcare System, Boston, MA, ^63^Center of Excellence for Stress & Mental Health, VA San Diego Healthcare System, San Diego, CA, 92161, USA, ^64^Center for Behavioral Genetics of Aging, University of California San Diego, La Jolla, CA, 92093, USA, ^65^Case Western Reserve University, Cleveland, OH, 44106, USA, ^66^Louis Stokes Cleveland VA Medical Center, Cleveland, OH, 44106, USA, ^67^VA Portland Health Care System, Portland, OR, 97239, USA

**VA Million Veteran Program COVID-19 Science Initiative Membership & Acknowledgements**

**VA Million Veteran Program COVID-19 Science Initiative**

**MVP COVID-19 Science Program Steering Committee**

- Christopher J. O’Donnell, M.D., M.P.H. (Co-Chair)

VA Boston Healthcare System, 150 S. Huntington Avenue, Boston, MA 02130

- J. Michael Gaziano, M.D., M.P.H. (Co-Chair)

VA Boston Healthcare System, 150 S. Huntington Avenue, Boston, MA 02130

- Philip S. Tsao, Ph.D. (Co-Chair)

VA Palo Alto Health Care System, 3801 Miranda Avenue, Palo Alto, CA 94304

- Sumitra Muralidhar, Ph.D.

US Department of Veterans Affairs, 810 Vermont Avenue NW, Washington, DC 20420

- Jean Beckham, Ph.D.

Durham VA Medical Center, 508 Fulton Street, Durham, NC 27705

- Kyong-Mi Chang, M.D.

Philadelphia VA Medical Center, 3900 Woodland Avenue, Philadelphia, PA 19104

- Juan P. Casas, M.D., Ph.D.

VA Boston Healthcare System, 150 S. Huntington Avenue, Boston, MA 02130

- Kelly Cho, M.P.H., Ph.D.

VA Boston Healthcare System, 150 S. Huntington Avenue, Boston, MA 02130

- Saiju Pyarajan, Ph.D.

VA Boston Healthcare System, 150 S. Huntington Avenue, Boston, MA 02130

- Jennifer Huffman, Ph.D.

VA Boston Healthcare System, 150 S. Huntington Avenue, Boston, MA 02130

- Jennifer Moser, Ph.D.

US Department of Veterans Affairs, 810 Vermont Avenue NW, Washington, DC 20420

**MVP COVID-19 Science Program Steering Committee Support**

- Lauren Thomann, M.P.H. (P&P Committee Representative, Working Group Coordinator)

VA Boston Healthcare System, 150 S. Huntington Avenue, Boston, MA 02130

- Helene Garcon, M.D. (Program Coordinator, Working Group Coordinator)

VA Boston Healthcare System, 150 S. Huntington Avenue, Boston, MA 02130

- Nicole Kosik, M.P.H. (Working Group Coordinator)

VA Boston Healthcare System, 150 S. Huntington Avenue, Boston, MA 02130

**MVP COVID-19 Science Program Working Groups and Associated Chairs**

- COVID-19 Related PheWAS
  - Katherine Liao, M.D.

VA Boston Healthcare System, 150 S. Huntington Avenue, Boston, MA 02130

- Scott Damrauer, M.D.

Philadelphia VA Medical Center, 3900 Woodland Avenue, Philadelphia, PA 19104

- Disease Mechanisms
  - Richard Hauger, M.D.

VA San Diego Healthcare System, 3350 La Jolla Village Drive, San Diego, CA 92161

- Shiuh-Wen Luoh, M.D., Ph.D.

Portland VA Medical Center, 3710 SW U.S. Veterans Hospital Road, Portland, OR 97239

- Sudha Iyengar, Ph.D.

VA Northeast Ohio Healthcare System, 10701 East Boulevard, Cleveland, OH 44106

- Druggable Genome
  - Juan P. Casas, M.D., Ph.D.

VA Boston Healthcare System, 150 S. Huntington Avenue, Boston, MA 02130

- Genomics for Risk Prediction, PRS, and MR
  - Themistocles Assimes, M.D., Ph.D.

VA Palo Alto Health Care System, 3801 Miranda Avenue, Palo Alto, CA 94304

- Panagiotis Roussos, M.D., Ph.D.

James J. Peters VA Medical Center, [130 W Kingsbridge Rd, Bronx, NY 10468](https://www.bing.com/local?lid=YN873x12457663610017047750&id=YN873x12457663610017047750&q=Emergency+Dept%2c+James+J+Peters+VA+Hospital&name=Emergency+Dept%2c+James+J+Peters+VA+Hospital&cp=40.86751174926758%7e-73.9051284790039&ppois=40.86751174926758_-73.9051284790039_Emergency+Dept%2c+James+J+Peters+VA+Hospital)

- Robert Striker, M.D., Ph.D.

William S. Middleton Memorial Veterans Hospital, 2500 Overlook Terrace, Madison, WI 53705

- GWAS & Downstream Analysis
  - Jennifer Huffman, Ph.D.

VA Boston Healthcare System, 150 S. Huntington Avenue, Boston, MA 02130

- Yan Sun, Ph.D.

Atlanta VA Medical Center, 1670 Clairmont Road, Decatur, GA 30033

- Pharmacogenomics
  - Adriana Hung, M.D., M.P.H.

VA Tennessee Valley Healthcare System, 1310 24th Avenue, South Nashville, TN 37212

- Sony Tuteja, Pharm.D., M.S.

Philadelphia VA Medical Center, 3900 Woodland Avenue, Philadelphia, PA 19104

- VA COVID-19 Shared Data Resource – Scott L. DuVall, Ph.D.; Kristine E. Lynch, Ph.D.; Elise Gatsby, M.P.H.

VA Informatics and Computing Infrastructure (VINCI), VA Salt Lake City Health Care System, 500 Foothill Drive, Salt Lake City, UT 84148

- MVP COVID-19 Data Core – Kelly Cho, M.P.H., Ph.D.; Lauren Costa, M.P.H.; Anne Yuk-Lam Ho, M.P.H.; Rebecca Song, M.P.H.

VA Boston Healthcare System, 150 S. Huntington Avenue, Boston, MA 02130

**VA Million Veteran Program**

**MVP Executive Committee**

- Co-Chair: J. Michael Gaziano, M.D., M.P.H.

VA Boston Healthcare System, 150 S. Huntington Avenue, Boston, MA 02130

- Co-Chair: Sumitra Muralidhar, Ph.D.

US Department of Veterans Affairs, 810 Vermont Avenue NW, Washington, DC 20420

- Rachel Ramoni, D.M.D., Sc.D., Chief VA Research and Development Officer

US Department of Veterans Affairs, 810 Vermont Avenue NW, Washington, DC 20420

- Jean Beckham, Ph.D.

Durham VA Medical Center, 508 Fulton Street, Durham, NC 27705

- Kyong-Mi Chang, M.D.

Philadelphia VA Medical Center, 3900 Woodland Avenue, Philadelphia, PA 19104

- Christopher J. O’Donnell, M.D., M.P.H.

VA Boston Healthcare System, 150 S. Huntington Avenue, Boston, MA 02130

- Philip S. Tsao, Ph.D.

VA Palo Alto Health Care System, 3801 Miranda Avenue, Palo Alto, CA 94304

- James Breeling, M.D., Ex-Officio

US Department of Veterans Affairs, 810 Vermont Avenue NW, Washington, DC 20420

- Grant Huang, Ph.D., Ex-Officio

US Department of Veterans Affairs, 810 Vermont Avenue NW, Washington, DC 20420

- Juan P. Casas, M.D., Ph.D., Ex-Officio

VA Boston Healthcare System, 150 S. Huntington Avenue, Boston, MA 02130

**MVP Program Office**

- Sumitra Muralidhar, Ph.D.

US Department of Veterans Affairs, 810 Vermont Avenue NW, Washington, DC 20420

- Jennifer Moser, Ph.D.

US Department of Veterans Affairs, 810 Vermont Avenue NW, Washington, DC 20420

**MVP Recruitment/Enrollment**

- Recruitment/Enrollment Director/Deputy Director, Boston – Stacey B. Whitbourne, Ph.D.; Jessica V. Brewer, M.P.H.

VA Boston Healthcare System, 150 S. Huntington Avenue, Boston, MA 02130

- MVP Coordinating Centers
  - Clinical Epidemiology Research Center (CERC), West Haven – Mihaela Aslan, Ph.D.

West Haven VA Medical Center, 950 Campbell Avenue, West Haven, CT 06516

- Cooperative Studies Program Clinical Research Pharmacy Coordinating Center, Albuquerque – Todd Connor, Pharm.D.; Dean P. Argyres, B.S., M.S.

New Mexico VA Health Care System, 1501 San Pedro Drive SE, Albuquerque, NM 87108

- Genomics Coordinating Center, Palo Alto – Philip S. Tsao, Ph.D.

VA Palo Alto Health Care System, 3801 Miranda Avenue, Palo Alto, CA 94304

- MVP Boston Coordinating Center, Boston - J. Michael Gaziano, M.D., M.P.H.

VA Boston Healthcare System, 150 S. Huntington Avenue, Boston, MA 02130

- MVP Information Center, Canandaigua – Brady Stephens, M.S.

Canandaigua VA Medical Center, 400 Fort Hill Avenue, Canandaigua, NY 14424

- VA Central Biorepository, Boston – Mary T. Brophy M.D., M.P.H.; Donald E. Humphries, Ph.D.; Luis E. Selva, Ph.D.

VA Boston Healthcare System, 150 S. Huntington Avenue, Boston, MA 02130

- MVP Informatics, Boston – Nhan Do, M.D.; Shahpoor (Alex) Shayan, M.S.

VA Boston Healthcare System, 150 S. Huntington Avenue, Boston, MA 02130

- MVP Data Operations/Analytics, Boston – Kelly Cho, M.P.H., Ph.D.

VA Boston Healthcare System, 150 S. Huntington Avenue, Boston, MA 02130

- Director of Regulatory Affairs – Lori Churby, B.S.

VA Palo Alto Health Care System, 3801 Miranda Avenue, Palo Alto, CA 94304

**MVP Science**

- Science Operations – Christopher J. O’Donnell, M.D., M.P.H.

VA Boston Healthcare System, 150 S. Huntington Avenue, Boston, MA 02130

- Genomics Core - Christopher J. O’Donnell, M.D., M.P.H.

VA Boston Healthcare System, 150 S. Huntington Avenue, Boston, MA 02130

Saiju Pyarajan Ph.D.

VA Boston Healthcare System, 150 S. Huntington Avenue, Boston, MA 02130

Philip S. Tsao, Ph.D.

VA Palo Alto Health Care System, 3801 Miranda Avenue, Palo Alto, CA 94304

- Data Core - Kelly Cho, M.P.H, Ph.D.

VA Boston Healthcare System, 150 S. Huntington Avenue, Boston, MA 02130

- VA Informatics and Computing Infrastructure (VINCI) – Scott L. DuVall, Ph.D.

VA Salt Lake City Health Care System, 500 Foothill Drive, Salt Lake City, UT 84148

- Data and Computational Sciences – Saiju Pyarajan, Ph.D.

VA Boston Healthcare System, 150 S. Huntington Avenue, Boston, MA 02130

- Statistical Genetics – Elizabeth Hauser, Ph.D.

Durham VA Medical Center, 508 Fulton Street, Durham, NC 27705

Yan Sun, Ph.D.

Atlanta VA Medical Center, 1670 Clairmont Road, Decatur, GA 30033

Hongyu Zhao, Ph.D.

West Haven VA Medical Center, 950 Campbell Avenue, West Haven, CT 06516

**Current MVP Local Site Investigators**

- Atlanta VA Medical Center (Peter Wilson, M.D.)

1670 Clairmont Road, Decatur, GA 30033

- Bay Pines VA Healthcare System (Rachel McArdle, Ph.D.)

10,000 Bay Pines Blvd Bay Pines, FL 33744

- Birmingham VA Medical Center (Louis Dellitalia, M.D.)

700 S. 19th Street, Birmingham AL 35233

- Central Western Massachusetts Healthcare System (Kristin Mattocks, Ph.D., M.P.H.)

421 North Main Street, Leeds, MA 01053

- Cincinnati VA Medical Center (John Harley, M.D., Ph.D.)

3200 Vine Street, Cincinnati, OH 45220

- Clement J. Zablocki VA Medical Center (Jeffrey Whittle, M.D., M.P.H.)

5000 West National Avenue, Milwaukee, WI 53295

- VA Northeast Ohio Healthcare System (Frank Jacono, M.D.)

10701 East Boulevard, Cleveland, OH 44106

- Durham VA Medical Center (Jean Beckham, Ph.D.)

508 Fulton Street, Durham, NC 27705

- Edith Nourse Rogers Memorial Veterans Hospital (John Wells., Ph.D.)

200 Springs Road, Bedford, MA 01730

- Edward Hines, Jr. VA Medical Center (Salvador Gutierrez, M.D.)

5000 South 5th Avenue, Hines, IL 60141

- Veterans Health Care System of the Ozarks (Gretchen Gibson, D.D.S., M.P.H.)

1100 North College Avenue, Fayetteville, AR 72703

- Fargo VA Health Care System (Kimberly Hammer, Ph.D.)

2101 N. Elm, Fargo, ND 58102

- VA Health Care Upstate New York (Laurence Kaminsky, Ph.D.)

113 Holland Avenue, Albany, NY 12208

- New Mexico VA Health Care System (Gerardo Villareal, M.D.)

1501 San Pedro Drive, S.E. Albuquerque, NM 87108

- VA Boston Healthcare System (Scott Kinlay, M.B.B.S., Ph.D.)

150 S. Huntington Avenue, Boston, MA 02130

- VA Western New York Healthcare System (Junzhe Xu, M.D.)

3495 Bailey Avenue, Buffalo, NY 14215-1199

- Ralph H. Johnson VA Medical Center (Mark Hamner, M.D.)

109 Bee Street, Mental Health Research, Charleston, SC 29401

- Columbia VA Health Care System (Roy Mathew, M.D.)

6439 Garners Ferry Road, Columbia, SC 29209

- VA North Texas Health Care System (Sujata Bhushan, M.D.)

4500 S. Lancaster Road, Dallas, TX 75216

- Hampton VA Medical Center (Pran Iruvanti, D.O., Ph.D.)

100 Emancipation Drive, Hampton, VA 23667

- Richmond VA Medical Center (Michael Godschalk, M.D.)

1201 Broad Rock Blvd., Richmond, VA 23249

- Iowa City VA Health Care System (Zuhair Ballas, M.D.)

601 Highway 6 West, Iowa City, IA 52246-2208

- Eastern Oklahoma VA Health Care System (Douglas Ivins, M.D.)

1011 Honor Heights Drive, Muskogee, OK 74401

- James A. Haley Veterans’ Hospital (Stephen Mastorides, M.D.)

13000 Bruce B. Downs Blvd, Tampa, FL 33612

- James H. Quillen VA Medical Center (Jonathan Moorman, M.D., Ph.D.)

Corner of Lamont & Veterans Way, Mountain Home, TN 37684

- John D. Dingell VA Medical Center (Saib Gappy, M.D.)

4646 John R Street, Detroit, MI 48201

- Louisville VA Medical Center (Jon Klein, M.D., Ph.D.)

800 Zorn Avenue, Louisville, KY 40206

- Manchester VA Medical Center (Nora Ratcliffe, M.D.)

718 Smyth Road, Manchester, NH 03104

- Miami VA Health Care System (Hermes Florez, M.D., Ph.D.)

1201 NW 16th Street, 11 GRC, Miami FL 33125

- Michael E. DeBakey VA Medical Center (Olaoluwa Okusaga, M.D.)

2002 Holcombe Blvd, Houston, TX 77030

- Minneapolis VA Health Care System (Maureen Murdoch, M.D., M.P.H.)

One Veterans Drive, Minneapolis, MN 55417

- N. FL/S. GA Veterans Health System (Peruvemba Sriram, M.D.)

1601 SW Archer Road, Gainesville, FL 32608

- Northport VA Medical Center (Shing Shing Yeh, Ph.D., M.D.)

79 Middleville Road, Northport, NY 11768

- Overton Brooks VA Medical Center (Neeraj Tandon, M.D.)

510 East Stoner Ave, Shreveport, LA 71101

- Philadelphia VA Medical Center (Darshana Jhala, M.D.)

3900 Woodland Avenue, Philadelphia, PA 19104

- Phoenix VA Health Care System (Samuel Aguayo, M.D.)

650 E. Indian School Road, Phoenix, AZ 85012

- Portland VA Medical Center (David Cohen, M.D.)

3710 SW U.S. Veterans Hospital Road, Portland, OR 97239

- Providence VA Medical Center (Satish Sharma, M.D.)

830 Chalkstone Avenue, Providence, RI 02908

- Richard Roudebush VA Medical Center (Suthat Liangpunsakul, M.D., M.P.H.)

1481 West 10th Street, Indianapolis, IN 46202

- Salem VA Medical Center (Kris Ann Oursler, M.D.)

1970 Roanoke Blvd, Salem, VA 24153

- San Francisco VA Health Care System (Mary Whooley, M.D.)

4150 Clement Street, San Francisco, CA 94121

- South Texas Veterans Health Care System (Sunil Ahuja, M.D.)

7400 Merton Minter Boulevard, San Antonio, TX 78229

- Southeast Louisiana Veterans Health Care System (Joseph Constans, Ph.D.)

2400 Canal Street, New Orleans, LA 70119

- Southern Arizona VA Health Care System (Paul Meyer, M.D., Ph.D.)

3601 S 6th Avenue, Tucson, AZ 85723

- Sioux Falls VA Health Care System (Jennifer Greco, M.D.)

2501 W 22nd Street, Sioux Falls, SD 57105

- St. Louis VA Health Care System (Michael Rauchman, M.D.)

915 North Grand Blvd, St. Louis, MO 63106

- Syracuse VA Medical Center (Richard Servatius, Ph.D.)

800 Irving Avenue, Syracuse, NY 13210

- VA Eastern Kansas Health Care System (Melinda Gaddy, Ph.D.)

4101 S 4th Street Trafficway, Leavenworth, KS 66048

- VA Greater Los Angeles Health Care System (Agnes Wallbom, M.D., M.S.)

11301 Wilshire Blvd, Los Angeles, CA 90073

- VA Long Beach Healthcare System (Timothy Morgan, M.D.)

5901 East 7th Street Long Beach, CA 90822

- VA Maine Healthcare System (Todd Stapley, D.O.)

1 VA Center, Augusta, ME 04330

- VA New York Harbor Healthcare System (Scott Sherman, M.D., M.P.H.)

423 East 23rd Street, New York, NY 10010

- VA Pacific Islands Health Care System (George Ross, M.D.)

459 Patterson Rd, Honolulu, HI 96819

- VA Palo Alto Health Care System (Philip Tsao, Ph.D.)

3801 Miranda Avenue, Palo Alto, CA 94304-1290

- VA Pittsburgh Health Care System (Patrick Strollo, Jr., M.D.)

University Drive, Pittsburgh, PA 15240

- VA Puget Sound Health Care System (Edward Boyko, M.D.)

1660 S. Columbian Way, Seattle, WA 98108-1597

- VA Salt Lake City Health Care System (Laurence Meyer, M.D., Ph.D.)

500 Foothill Drive, Salt Lake City, UT 84148

- VA San Diego Healthcare System (Samir Gupta, M.D., M.S.C.S.)

3350 La Jolla Village Drive, San Diego, CA 92161

- VA Sierra Nevada Health Care System (Mostaqul Huq, Pharm.D., Ph.D.)

975 Kirman Avenue, Reno, NV 89502

- VA Southern Nevada Healthcare System (Joseph Fayad, M.D.)

6900 North Pecos Road, North Las Vegas, NV 89086

- VA Tennessee Valley Healthcare System (Adriana Hung, M.D., M.P.H.)

1310 24th Avenue, South Nashville, TN 37212

- Washington DC VA Medical Center (Jack Lichy, M.D., Ph.D.)

50 Irving St, Washington, D. C. 20422

- W.G. (Bill) Hefner VA Medical Center (Robin Hurley, M.D.)

1601 Brenner Ave, Salisbury, NC 28144

- White River Junction VA Medical Center (Brooks Robey, M.D.)

163 Veterans Drive, White River Junction, VT 05009

- William S. Middleton Memorial Veterans Hospital (Robert Striker, M.D., Ph.D.)

2500 Overlook Terrace, Madison, WI 53705

**Supplemental Methods**

**Data Sources**

In this COVID-19 study, we used data from MVP, a large multi-ethnic genetic biobank at Veterans Affairs. MVP began enrolling veterans in 2011 and has now enrolled over 850,000 veterans and genotyped over 650,000. The Veterans' EHR data included diagnosis codes (ICD-9 and ICD-10), current procedural terminology codes (CPT), clinical laboratory measures, and demographic, lifestyle, and radiology reports (44). Pre-COVID EHR data were collected from participants from the time of enrollment to September 30, 2019.

The MVP cohort SNP data was generated using a custom Thermo Fisher Axiom genotyping platform named MVP 1.0. The MVP cohort quality control and genotyping imputation steps were previously reported(45). Ancestry of participants was defined using Harmonized ancestry, race, and ethnicity (HARE), a variable derived from self-reported survey data and genetically derived ancestry(15). Directly genotyped rs35705950-T information was extracted and utilized for association testing.

A VA-wide collective effort lead by the VA Informatics and Computing Infrastructure (VINCI) created a shared COVID-19 Shared Data Resource (SDR) during the April-August 2020 period, providing a new data domain related to COVID-19 for the VA. All metadata on documentation regarding conditions, laboratory measures, medications, and procedures pertaining to the COVID-19 pandemic and dissemination of information regarding SDR were provided by the VA Phenomics Library, Centralized Interactive Phenomics Resource (CIPHER), to users across the VA healthcare systems. The VA COVID-19 SDR group including the VA National Surveillance Team provided researchers with curated data extracted from electronic health records (EHR) of Veterans, after stripping off identifiable elements, and ensuring data security by requiring restricted access to the data(41). This activity was undertaken centrally by an experienced VA research team in order to ensure consistent applications of dates across the EHR tables, as well as uniform definitions of events preceding and subsequent to COVID-19, in an effort to characterize the trajectory of COVID-19 and other diseases and conditions. VA-wide efforts describing these data are already in public domain(42).   Structured data obtained via CPT and ICD9 or ICD10 codes, deposited in the VA Corporate Data Warehouse (CDW),was further enriched with rule-based unstructured events recorded in patient notes via natural language processing (NLP), as was done previously for other projects(43).  The purpose of NLP-boosting was to fill gaps in knowledge about the severity of the disease, as well as extract specific dates when procedures (e.g. intubation and extubation) were performed. These curated data were then provisioned in the Million Veteran Program (MVP) study mart for COVID-19 in VINCI ensuring data security behind the VA firewall.

*COVID-19 outcome definitions*

The primary analysis *MUC5B* SNP rs35705950-T with COVID-19 infection and hospitalization included all the patients who were tested from Feb 2020 through April 2021 using RT-PCR based method(16, 17) or self-reported during primary care visit.  These patients may have received their COVID-19 PCR testing inside as well as outside of the VA.  However, for analysis of the outcome severity of COVID-19 infection such as ICU stay or death (more details below), we only included patients tested within VA. The *index date* was defined as a COVID-19 diagnosis date, i.e., specimen date, or a self-reported date of diagnosis; and for a hospitalized patient, the admission date up to 15 days prior to the COVID-19 case date.  “*Pre-index*” and “*post-index*” are hereafter used to denote events and outcomes before and after this *index date*. To define the COVID-19 related severity and death, we use a modified version of COVID-19 Severity Scale variable from WHO COVID-19 Disease Progression Scale(18).  The current version of VA’s adaptation of COVID-19 Severity Index classifies severity into mild, moderate, severe and death. The severity rating of COVID-19 infection for the MVP cohort was modified from the WHO criteria to classify into four groups: mild (testing positive for SARS-CoV-2 by PCR); moderate (SARS-CoV-2 positivity and hospitalization within 30 days of index dates); severe (SARS-CoV-2 positivity, hospitalized and  having indication of treatment by noninvasive mechanical ventilation (NIV), high flow oxygen support, extracorporeal membrane oxygenation (ECMO), intubation, vasopressor support, and dialysis; and death within 30 days of index dates for COVID-19 infection (18)). All data and variables were provided by the MVP data core.

We applied two approaches to define COVID-19 outcomes. First, for the meta-analysis with HGI summary statistics, we synchronized the phenotype definitions as used by the consortium. The following binary COVID-19 outcomes were examined: (1) COVID Susceptibility: COVID-19 positive vs. population controls; (2) COVID Hospitalization-v1: COVID-19 positive and hospitalized vs. population controls; (3) COVID-19 hospitalization-v2: COVID-19 positive and hospitalized vs. COVID-19 positive but not hospitalized. Secondly, we defined two other binary outcomes using patients who received COVID-19 PCR testing at VA sites: 4) severe COVID-19 (COVID19 severe cases based on WHO criteria described above vs. testing positive for COVID-19 but not severe); 5) death (death within 30 days of COVID-19 infection vs. all other COVID-19 test positives).

*Phenotyping of pre- and post-index conditions for COVID-19*

To determine pre-index conditions, we used NLP-boosted unstructured notes, ICD and CPT codes, and medications taken 2 years prior. ICD and CPT codes and medications 60 days after the index date were used to derive post-index conditions. Pre- and post-index conditions were obtained from the Shared Data Resource's pre- and post-index tables (SDR). The study's post-index conditions' ICD codes are in the supplemental materials. Post-index pneumonia (Pneumonia60d) was studied 60 days after COVID-19 index dates (**Table E2**).

For the 60-day post-index analysis, the SDR post-index tables were used. We included only patients whose PCR testing was done at the VA. Additionally patients that were initially tested negative in the Patient table but then turned positive in the post-index table were excluded from the post index analysis. Participants who lost follow up in the post-index table were also excluded.

The cohort demographics and clinical conditions documented for all tested patients within the two years preceding the index dates are presented in a supplemental table (**Table E1**).

**Meta-analysis with HGI**

Results from the COVID-19 Host Genetics Initiative (HGI)(46) were utilized for replication and meta-analysis with our results. Since MVP contributed to HGI, we used the “leave one out” results that were available only to contributing cohorts. These are currently available only for Release 5 (available January 18, 2021), not the more recent Release 6 (June 15, 2021), and also excluded 23&Me, per their agreement with HGI. Results for rs35705950-T were extracted for both the European-only and trans-ethnic meta-analysis across all phenotype definitions.

 Logistic regression was performed to determine the association of rs35705950-T with each of these outcomes within each of three HARE AFR, HIS, and EUR ancestry) within plink2a(24) adjusting for age, sex, sex^2^, and the first 15 ethnicity-specific principal components. Inverse-variance weighted meta-analysis was performed in GWAMA(25) to combine MVP with HGI.

**Phenome-wide association study (PheWAS)**

For the PheWAS analysis on the *MUC5B* rs35705950-T allele, we used Phecodes (23) and laboratory measures from the clinical data available prior to the onset of COVID-19 infection (Sept 2019). Individuals with two or more Phecodes were defined as cases, while those without were defined as controls. In our previous study, we found that cases under 200 provide >80% statistical power and low type I error therefore we excluded phecodes with fewer than 200 cases within each ancestry group. It yielded 1618 (EUR), 1289 (AFR), 994 (HIS) Phecodes. We also performed a laboratory-wide association study using 69 clinical labs extracted from the EHR of MVP participants.  We used the median of each person's entire lab history. We used logistic regression for Phecodes and linear regression for laboratory measurements in PLINK2(24). When the logistic regression model failed to converge for binary outcomes, firth regression was used. Regression models were adjusted for sex, age (at the time of data freeze), quadratic term of age, and the first 20 principal components. We applied Bonferroni correction to each ancestry specific analysis to adjust for multiple hypothesis testing, and following threshold were use select significant associations: EUR = 3.09 x 10^-05^ (0.05/1618), AFR = 3.8 x 10^-05^ (0.05/1289), EUR = 5.03 x 10^-05^(0.05/994).

46.	Home. at <<https://www.covid19hg.org/>>.

**Supplemental Figure**

**Figure E1.** Overview of genetic association of MUC5B allele and COVID-19 phenotypes as well as phenome-wide association study.

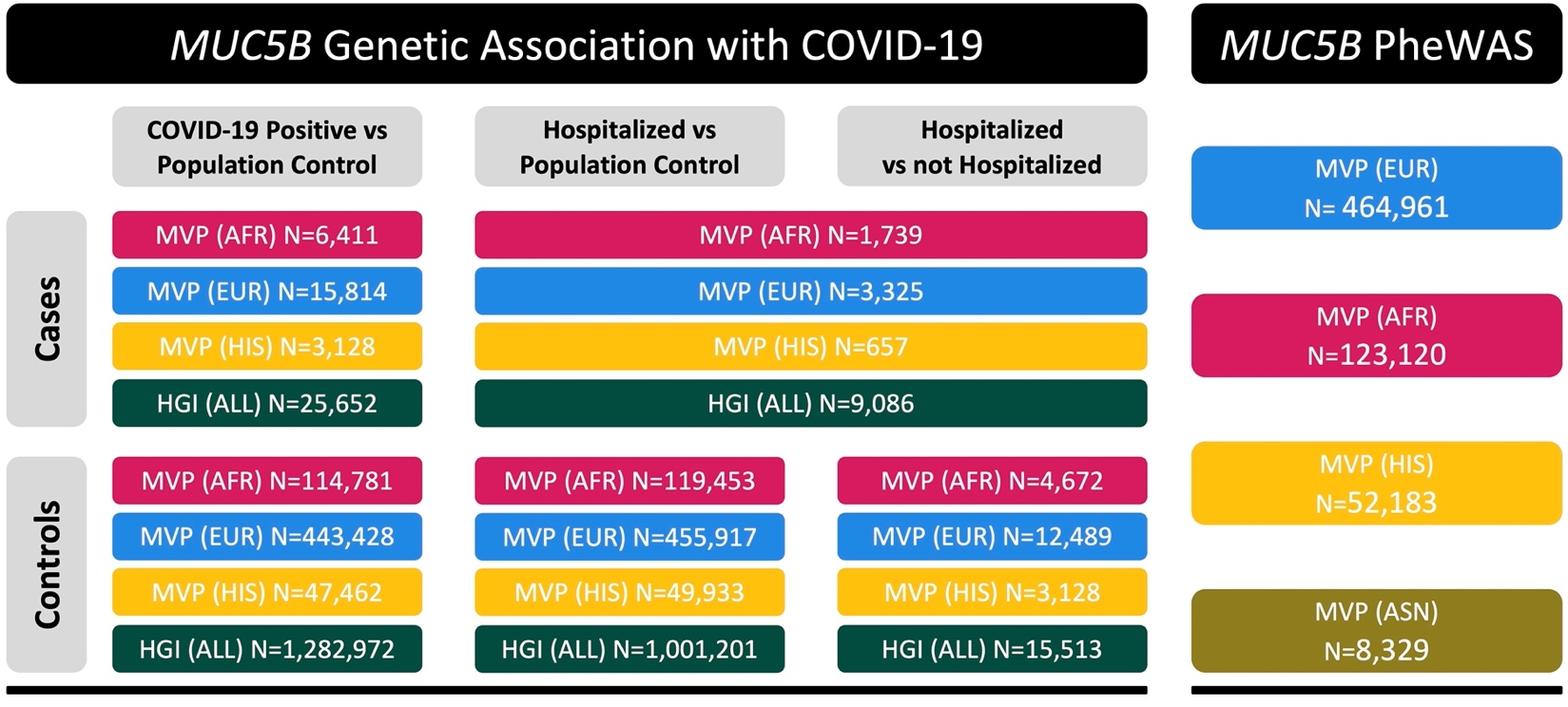

**Supplemental Tables**

**Table E1**. The demographics and clinical conditions documented within two years prior to the index dates for MVP subjects used for association studies with severe outcomes of COVID-19 infection and post-index pneumonia events (pneumonia60d) in this study.

**See Excel Document**

**Table E2.** The ICD codes used for data pull of post-index pneumonia diagnosis (pneumonia60d) in this study.

**See Excel Document**

**Table E3**. Full summary statistics of PheWAS associations with *MUC5B* rs35705950 allele.

**See Excel Document**

**Table E4.** LabWAS reveals disease conditions associated with the presence of a *MUC5B* rs35705950 allele.

**See Excel Document**

**Table E5.** Sample sizes and rates of pneumonia within 60 days post index date for MVP subjects with European ancestry with no missing for pre- or post- index pneumonia.

|  | **rs35705950 _0 copy** | | **rs35705950 _1 copy** | | **rs35705950 _2 copies** | |
| --- | --- | --- | --- | --- | --- | --- |
| HARE | Pneumonia_no | Pneumonia_yes | Pneumonia_no | Pneumonia_yes | Pneumonia_no | Pneumonia_yes |
| **COVID-19 Tested Positive** | | | | | | |
| AFR | 26948 | 775 | 1141 | 24 | 18 | -- |
| EUR | 62451 | 2117 | 15186 | 537 | 928 | 31 |
| HIS | 9687 | 249 | 1574 | 32 | 65 | 1 |
| **COVID-19 Tested Negative** | | | | | | |
| AFR | 2800 | 966 | 125 | 32 | 1 | 1 |
| EUR | 5185 | 1490 | 1262 | 310 | 73 | 15 |
| HIS | 1309 | 366 | 187 | 68 | 9 | 1 |

**Table E6.** Association of rs35705950 allele with two adverse outcomes of COVID-19 1) Severe COVID-19 + Death and 2) COVID-19 related Death

|  | **rs35705950 _0 copy** | | **rs35705950 _1 copy** | | **rs35705950 _2 copies** | |  |
| --- | --- | --- | --- | --- | --- | --- | --- |
| **Severe COVID-19 + Death** | | | | | | | |
| HARE | Severe plus death_no | Severe plus death_yes | Severe plus death_no | Severe plus death_yes | Severe plus death_no | Severe plus death_yes | Odds ratio  [95% CI] |
| AFR | 3693 | 440 | 155 | 16 | 2 | 0 | 0.87 [0.51, 1.47] |
| EUR | 6740 | 636 | 1593 | 150 | 91 | 6 | 1.01 [0.85, 1.20] |
| HIS | 1649 | 165 | 252 | 18 | 9 | 1 | 0.76 [0.48, 1.23] |
| **COVID-19 related Death** | | | | | | | |
| HARE | Death_no | Death_yes | Death_no | Death_yes | Death_no | Death_yes | Odds ratio [95% CI] |
| AFR | 3953 | 180 | 161 | 10 | 2 | 0 | 1.48 [0.77, 2.84] |
| EUR | 7031 | 345 | 1671 | 72 | 94 | 3 | 0.91 [0.72, 1.16] |
| HIS | 1734 | 80 | 263 | 7 | 9 | 1 | 0.75 [0.39, 1.45] |
